# Attitude and subjective norms as predictors of intention of married men and women to accept caesarean section delivery in Lagos, Nigeria

**DOI:** 10.1101/2025.04.24.25326363

**Authors:** Titilayo Aike Olaoye, Boluwatife Adebambo, Blessing Osie-Efetie, Ololade Ogunsanmi, Clarita Panuel-Egwakhe, Saratu Ajike&, Juliana Olwatoyin Elebiju, Favour Ayobola Ojo, Nnodimele Onuigbo. Atulomah

**Affiliations:** Department of Public Health, Babcock University Ilishan-Remo Ogun state, Nigeria; Department of Health and Safety education, Delta State University, Abraka, Delta State, Nigeria; Department of Public Health, Adeleke University Ede, Osun State Nigeria

**Keywords:** Attitude, Caesarean Section, Couples, Intention to Accept, Subjective norms

## Abstract

**Background:** Despite the life-saving benefits, there is a great deal of reluctance among Nigerian women and their husbands to have a caesarean section (CS) delivery for at risk pregnancy. This study assessed attitude and associated subjective norm involved in resolving whether to elect CS delivery among couples in Lagos, Nigeria.

**Methods:** The study was cross-sectional survey design. A validated questionnaire was used to collect data from four hundred and twenty-two consenting respondents who were selected through a systematic sampling technique. Data were transformed from response items into weighted-aggregate scores for moderating variables. The statistical product and Service Solution version 23 was used in analyzing the data to generate mean, standard deviation and multivariate analysis. All test were conducted at p ≤ 0.05 level of significance.

**Results:** The mean age of the respondents for males and females were 38.7±11.6 years and 37.4±9.8 years respectively. The male and female respondents had a mean score of 7.78 ±2.82; 8.56 ±3.08; 15.41±6.97 and 13.66 ± 7.10 computed for attitudinal disposition and subjective norms respectively. Three out of every 10 male respondents and 2 out of every 10 females were likely to accept CS if the need arose. Pain accompanied with the delivery method (male: 40.3%; female: 38.4%) and risk/complications of the method (male: 30.3%; female: 23.7%) were barriers the respondents reported. Over half of the respondents (males: 53.1%; females: (57.8%) had moderate level of intention to accept CS. There was a significant association between the subjective norm considerations of both the male (R^2^ = 0.04; Beta= 0.08; *p* = 0.002) and female (R^2^ = 0.09; Beta= 0.12; *p* = 0.0001) respondents regarding their intention to accept CS.

**Conclusion:** Most of the respondents placed high subjective norm considerations in accepting CS. None of the respondents expressed high intention of accepting CS. Alongside efforts to strengthen health education regarding CS option for delivery, the media should be used as a tool to debunk misconceptions about CS actively.

## Introduction

Childbirth is the process through which one or more babies exit the uterus via vaginal delivery or caesarean section^1^. It is a transformative and life-changing experience for both women and their families, and the choice of delivery method plays a pivotal role in the birth process. While vagina delivery remains the most common form of childbirth globally^2^, the rate of caesarean-section (CS) deliveries has particularly increased in developed countries^3^. This increase has been associated with factors such as maternal age, medical advancement and evolving obstetric practices^3^. A cesarean section, an obstetric procedure that involves surgical delivery of a baby through making an incision in the woman’s abdominal wall and uterus^4^, is one of the components of comprehensive emergency obstetric care intervention provided for pregnant women to promote safe delivery^5^.

Caesarean section is not only safe but is also medically advisable in certain instances. To promote the survival and well-being of the women and their newborns, women with high-risk pregnancies like multiple fetuses, breech presentation, uterine rupture, obstructed labour and those with transmissible infection are usually advised to deliver through C-section^6, 7^. The importance of caesarean sections in preventing difficulties during childbirth and lowering maternal and foetal mortality rates is widely recognised^8^. Medical professionals also agree that although it has a higher chance of complications than a vaginal delivery^9^; a caesarean section (CS) is safe.

Annually, over 18.5 million caesarean section deliveries are registered globally, accounting for 19.1% of overall childbirths on average, with regional variations^8, 10^. It has been projected that by 2030, 28.5% of women worldwide will give birth through C-section (38 million caesareans annually, of which 33.5 million will occur in LMIC) ^11^. In 2015, the World Health Organization recommended that the optimum caesarean section rate at the population level should be between 10% and 15%. There is an increased rate of caesarean delivery in countries such as the United States, Australia, Brazil and China where many of these C-section deliveries are unnecessary and medically unjustifiable^12^. The most recent caesarean delivery prevalence rate reported in Nigeria was 2.7% and despite the high fertility, maternal and perinatal mortality rate in Nigeria; this is low compared with the recommendation of the World Health Organization^13^. This low prevalence suggests unmet needs which reflect the high maternal and perinatal mortality rate experienced in the country^13^, also owing to poor acceptability of C-section, limited access to the service, and health-system inadequacies^13, 14^.

The acceptability of C-section depends on personal and cultural views, regardless of improvements in C-section technique, safe anaesthesia, upgraded or more effective antibiotics, provision of blood transfusion services, and more education and enlightenment^15^. In many developing nations such as Nigeria, pessimistic views continue to exist among a lot of women and their families regarding C-section delivery methods. Women who have had caesarean deliveries are seen as weaklings and reproductive failures in most of these societies, and some are even said to be the result of an unfaithful wife’s curse^16^. Other reasons include high mortality rate, extended hospital stays, and the perceived high expense of hospital bills^15^.

In addition, anecdotal reports in Lagos State suggest that some pregnant women refused to accept caesarean section delivery, even when advised by health workers (nurses/doctors) to deliver their babies through caesarean section at no cost. However, they rather go to prayer houses or traditional birth attendants, where some of them experience prolonged labour, preventable complications and even the death of their baby.

Similarly, women in Ghana perceived CS delivery as “troublesome and rendering them unable to perform domestic and economic duties that were detrimental to their families’ survival” ^17^.These beliefs, if widely accepted, have the potential to reduce reliance on the medically necessary C-section, thereby increasing neonatal and perinatal mortality, particularly among rural women^17^.

The Theory of Reasoned and Action (TRA) explains that an individual’s intention is the predictor of the behaviour. This theory posits that individual behaviour is influenced by two main factors: attitude and subjective norm, which in turn are triggered by the individual’s belief and the perception of others. In the context of this study, the intention, of both married men and women to accept C-section delivery, is shaped by the combination of their attitudes towards the procedure which are influenced by an individual’s belief about the consequences of performing the behaviour and subjective norms (individual perception of social pressure to perform or not perform C-section) (See figure 1).

**Figure 1:**
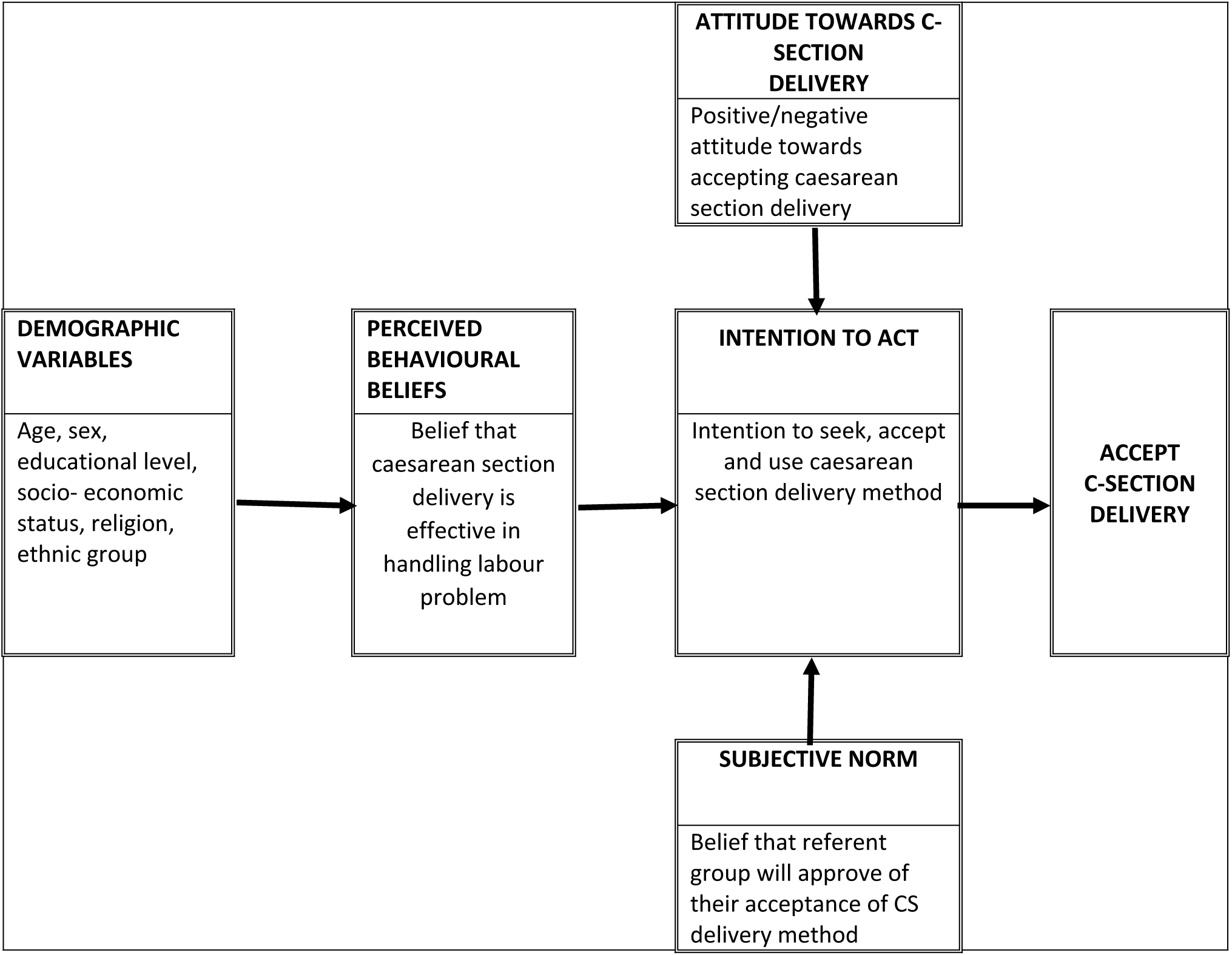
Conceptual framework derived from Theory of Reasoned Action guiding the study.

Investigating the interaction between these factors could inform policymakers, program planners and healthcare professionals to plan interventions that can support couples in making informed decisions regarding childbirth, thereby reducing maternal and neonatal mortality. Also, this study will contribute to a broader understanding of how subjective norm influences interact with reproductive health behaviour of individuals in Lagos State.

Most studies conducted on caesarean sections focused mainly on beliefs, perceptions of women, and social-cultural barriers to acceptance of CS^15, 18.19^. There is a need to include the male partners who have a major role in child birth decision-making. Hence, this study utilized the Theory of Reasoned Action (TRA) in investigating attitude and associated subjective norm involved in resolving whether to elect CS delivery among couples in Lagos, Nigeria.

## Methods

### Research design

This study employed a cross-sectional survey design that collected data from consenting participants selected from six communities in Ikorodu Local Government Area.

### Study Location

This study took place in Ikorodu Local Government Area (LGA). Ikorodu lies 36 Kilometres North of Lagos and is bordered on the South by the Lagos Lagoon, on the North by a border with Ogun State, and on the East by a border with Agbowa-Ikosi. In 2003 the exiting Ikorodu LGAs was split for administrative purposes into Local Council Development Areas (LCDAs). These lower-tier administrative units are now six (6): Imota, Igbogbo/Bayeku, Ijede, Ikorodu North, Ikorodu West and Ikorodu Central with an estimated population of 527,917 comprising of 268,463 males and 259,454 females^20^.

### Inclusion and exclusion criteria

Eligible respondents were married men and women who consented to participate in the study. Respondents, who were sick or not mentally stable, were excluded.

### Sample Size Determination

This study involved a total of 422 respondents drawn from six communities calculated by employing the formula used by Cochran (1965) in a previous study on the bases that the population were more than 10,000.

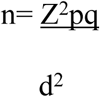

where,
Where n denotes the estimate of the sample size for the married men and women Z= 1.96 derived from 95% confidence level of the Standard normal distribution to account for Type I error.

p= Prevalence which is assumed to be 50% =0.5; q= 1-p= 0.5 d= Degree of accuracy which is at 5% =0.05 Therefore, sample size 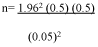

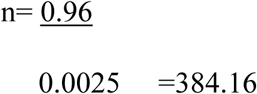

The study applied 10 % on the minimum sample size computed to account for non-response bias giving a final sample size of 422 participants.

The study participants were selected using multistage sampling procedure. In the first stage simple random sampling by balloting was used to select three Local Council Development Areas (LCDAs) out of the six LCDAs. The three selected LCDA were Ikorodu Central, Ikorodu North and Igbogbo-Baiyeku. The second stage involved the selection of 6 wards (2 wards per LCDA) from the selected LCDAs using simple random sampling by balloting. Furthermore, the third stage involved purposive selection of the largest community per ward yielding 6 communities. In the selection of the participants, the number of streets and building per streets were considered. One street was selected per community and a proportionate sampling technique was used to select participants per street. An average of 2 participants, who met the inclusion criteria, was selected per building through balloting.

## Variables

### Explanatory variables and dependent variables

The explanatory variables considered were the socio-demographic characteristics as independent variables while attitudinal disposition as moderating variable measured on a 15-point rating scale using the 4-point Likert-type response scales of (strongly agree, agree, strongly disagree and disagree, and the level of subjective norm consideration measured on a 4-point Likert-type response scales of (Strongly agree, agree, strongly disagree and disagree). The dependent variable was intention of accepting CS delivery. It assessed the participants’ readiness to use CS delivery when the need arises. The dependent variable consisted of three items measured on 3-Point likert-type response scales of Likely, unlikely and Not at all; the least being scored a zero point and the highest a 6-point. Hence, intention to accept CS was measured on a 15-point rating scale. The response-items were transformed to weighted-aggregate scores to generate the overall scores to enable computation of summaries of descriptive statistics.

### Instrument validity and Reliability

The instrument was developed in English and translated into Yoruba because Ikorodu is home to the majority of Yoruba ethnic people (a major ethnic group of Nigeria living in the western states of the country). As a result, respondents could select the questionnaire in the language of their choice. The questionnaire was developed guided by the Theory of Reasoned Action (TRA) and provided the basis for construct validity of the instrument; also literatures provided validation for the relevance of the variables in the study. The instrument’s internal consistency was measured by pre-testing the instrument with 42 participants in Sagamu. A Cronbach’s alpha score of 0.78 was obtained after necessary corrections.

### Data collection Procedure

Three community influencers were recruited and trained as research assistants on the study processes. Iinformed consent (verbal) was obtained before administering the questionnaire and the study was also introduced to the participants by the research assistants. Further assistance was given to some participants who could not read.

### Data Analysis and test of significance

A coding guide was used to translate the responses obtained from the questionnaire. The attitudinal disposition, subjective norms and intention to accept CS data were transformed into a rating scale to derive standard measures. The IBM Statistical Package for Service Solution (IBM SPSS) version 29.0 was used for data analysis. Univariate descriptive analysis of socio-demographic, attitude, subjective norms and intention to accept caesarean section variables were performed. Also, Analysis of variance (ANOVA) was performed to test differences in the level of intention to accept caesarean section across the socio-demographic profiles. Linear regression analysis was conducted to establish the degree of association between the computed variable of intention to accept caesarean section with the TRA construct. Data were summarized and presented on frequency tables with percentages. Statistical tests were conducted at cut-off of p ≤0.05.

*Ethical Consideration*: Ethical approval to conduct the study was granted by Babcock University Health and Research Ethical Committee [BUHREC No: 959/21] after a thorough review of the study protocol to ensure the safety of the participants and their rights. Participants were briefed about the nature and extent of the study and verbal consent was given; no names were written in the questionnaires to ensure confidentiality and participation in the study was voluntary.

## RESULTS

### Socio-demographic characteristics

The study enrolled 422 consenting participants, comprising 212 males and 210 females. Most respondents (61.3% of males and 63.3% of females) were aged 20-40 years, with a mean (x-) ± SD of 38.67 ± 11.67 years for males and 37.46 ±9.88 years for females. Christianity was the predominant religion, with 51.9% of males and 57.6% of females. Yoruba individuals represented the largest ethnic group, comprising 63.7% of males and 62.4% of females. Regarding educational attainment, the highest proportion of males held a bachelor’s degree (31.1%), while more than a quarter of females (31.0%) had an HND/OND (Higher National Diploma/Ordinary National Diploma). Less than half of the respondents were employed (males = 43.4%, female = 39.5%). Upon assessment, the lowest mean scores on the intention to accept caesarean section delivery was found among male atheists (6.38), female followers of African Traditional Religion (5.92), Hausa males (6.23), Yoruba females (6.91), individuals with primary education (5.57 for males and 6.08 for females), unemployed males (6.25) and self-employed females (6.41) (See Table 1).

**Table 1:** Demographic characteristics of the respondents.

| **Variables** | **N (%)**<br>**Male (n=**<br>**212)** | **Mean scores**<br>**and 95%CI on**<br>**Intention to**<br>**accept**<br>**Caesarean**<br>**section (CS)**<br>**delivery** | **N (%)**<br>**Female**<br>**(n=210)** | **Mean scores and**<br>**95%CI on Intention to**<br>**accept Caesarean**<br>**section (CS) delivery** |
| --- | --- | --- | --- | --- |
| **Age (in years)** |  |  |  |  |
| **20-40** | 130(61.3) | 6.96(6.51-7.42) | 133 (63.3) | 7.20(6.76-7.65) |
| **40-60** | 71(33.5) | 7.46(6.80-8.03) | 74(35.2) | 6.61(5.96-7.31) |
| **60-90** | 11 (5.2) | 5.91(4.27-7.45) | 3(1.4) | 7.67(7.00-8.00) |
| **Religion** |  |  |  |  |
| **Christianity** | 110(51.9) | 7.30(6.78-7.77) | 121(57.6) | 7.07(6.05-9.39) |
| **Islam** | 86 40.6) | 6.88(6.27-7.42) | 75(35.7) | 7.04(6.43-7.62) |
| **African** | 8 (3.8) | 6.75(5.50-8.22) | 13(6.2) | 5.92(4.10-7.55) |
| **Traditional** | 8 (3.8) | 6.38(3.29-9.67) | 1(0.5) | 10.00(10.00-10.00) |
| **Religion** |  |  |  |  |
| **Atheist** |  |  |  |  |
| **Ethnicity** |  |  |  |  |
| **Igbo** | 51(24.1) | 7.35(6.53-8.02) | 60(28.6) | 6.93(6.27-7.52) |
| **Yoruba** | 135(63.7) | 7.17(6.70-7.53) | 131(62.4) | 6.91(6.42-7.34) |
| **Hausa** | 21(9.9) | 6.23(4.71-7.52) | 17(8.1) | 7.71(5.94-9.00) |
| **Others** | 5 (2.4) | 5.20(2.00-8.40) | 2(1.0) | 9.00(8.00-10.00) |
| **Level of** |  |  |  |  |
| **Education** | 14 (6.6) | 5.57(3.86-7.38) | 12(5.7) | 6.08(4.14-7.66) |
| **Primary school** | 31(14.6) | 5.97(4.57-16.38) | 56(26.7) | 6.77(5.98-7.47) |
| **SSCE** | 63(29.7) | 7.32(6.60-7.92) | 65(31.0) | 6.82(6.09-7.49) |
| **HND/OND** | 66 (31.1) | 7.38(6.71-8.01) | 52(24.8) | 7.31(6.53-8.04) |
| **BSc** | 33(15.6) | 7.52(6.75-8.26) | 23(11.0) | 7.96(7.28-8.71) |
| **MSc** | 5(2.36) | 8.20(7.00-10.00) | 2(1.0) | 6.00(3.00-9.00) |
| **PHD** |  |  |  |  |
| **Occupation** |  |  |  |  |
| **Unemployed** | 24(11.3) | 6.25(4.84-7.57) | 83(8.1) | 7.00(5.57-8.34) |
| **Employed** | 92 (43.4) | 7.62(6.71-7.77) | 17 (39.5) | 6.95(6.32-7.54) |
| **Self –** | 29(13.7) | 6.41(5.14-7.54) | 54(25.77) | 6.41(5.14-7.15) |
| **employed** | 18(8.5) | 7.39(6.39-8.42) | 19 (9.0) | 7.40(6.39-8.42) |
| **Artisan** | 49(23.1) | 7.41(6.67-8.06) | 37 (17.6) | 7.41(6.67-8.06) |
| **Civil servant** |  |  |  |  |

## TRA-constructs

### Attitudinal disposition towards Caesarean Section (CS)

The findings presented here highlight attitudes toward caesarean section (CS) birth. More than half of the male respondents (54.3%) and nearly half of the female respondents (47.6%) perceived CS as an unnatural way of childbirth. Likewise, approximately 7 in 10 males (68.9%) and 6 in 10 females (59.1%) believe that undergoing CS is a sign of weakness. Over half of the male respondents (54.3%) and a comparable proportion of females (57.2%) strongly believed that a woman should only have a CS with her husband’s approval. Additionally, nearly two-thirds of the male (66.5%) and female (65.2%) respondents perceived CS as profit making venture rather than a necessary medical intervention. Similarly, the belief that CS is unnecessary because childbirth should happen naturally is common among about 6 in 10 males (60.8%) and females (56.2%). Overall, there respondents had positive attitude towards CS (refer to Table 2 for details).

**Table 2:**
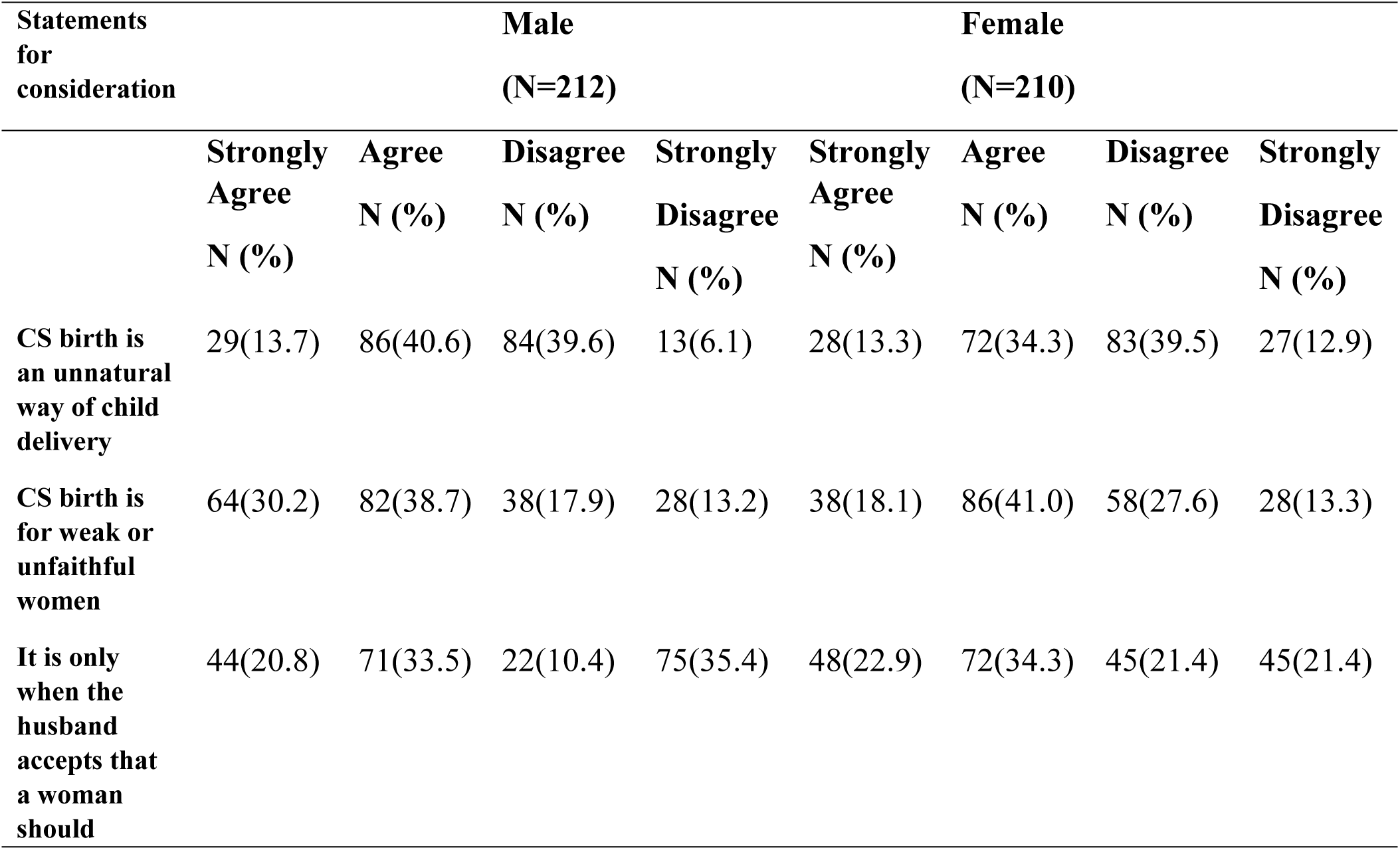
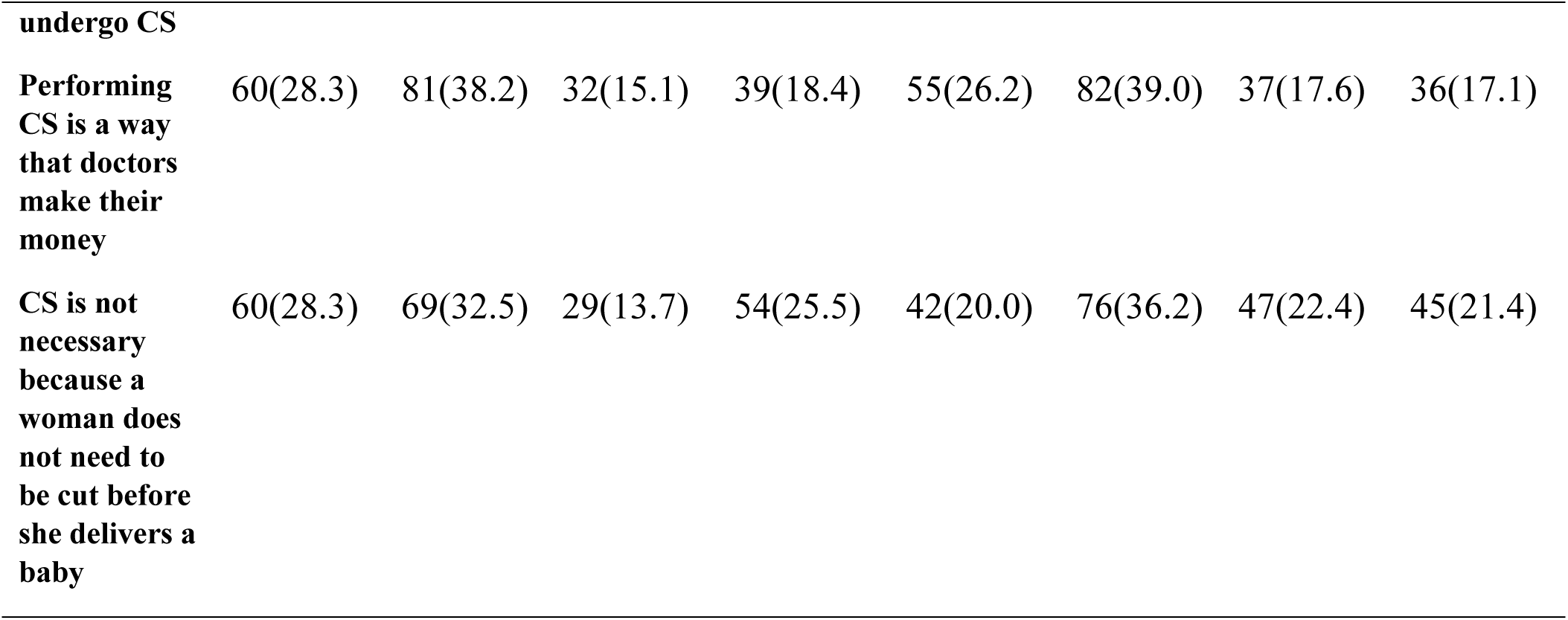
Frequency distribution of responses to items reflecting Attitudinal disposition towards CS.

| Statements<br>for<br>consideration |  | Male<br>(N=212) |  |  |  | Female<br>(N=210) |  |  |  |
| --- | --- | --- | --- | --- | --- | --- | --- | --- | --- |
|  |  | Strongly<br>Agree<br>N (%) | Agree<br>N (%) | Disagree<br>N (%) | Strongly<br>Disagree<br>N (%) | Strongly<br>Agree<br>N (%) | Agree<br>N (%) | Disagree<br>N (%) | Strongly<br>Disagree<br>N (%) |
| CS birth is<br>an unnatural<br>way of child<br>delivery |  | 29(13.7) | 86(40.6) | 84(39.6) | 13(6.1) | 28(13.3) | 72(34.3) | 83(39.5) | 27(12.9) |
| CS birth is<br>for weak or<br>unfaithful<br>women |  | 64(30.2) | 82(38.7) | 38(17.9) | 28(13.2) | 38(18.1) | 86(41.0) | 58(27.6) | 28(13.3) |
| It is only<br>when the<br>husband<br>accepts that<br>a woman<br>should |  | 44(20.8) | 71(33.5) | 22(10.4) | 75(35.4) | 48(22.9) | 72(34.3) | 45(21.4) | 45(21.4) |

|  |  |  |  |  |  |  |  |  |
| --- | --- | --- | --- | --- | --- | --- | --- | --- |
| **undergo CS** |  |  |  |  |  |  |  |  |
| **Performing CS is a way that doctors make their money** | 60(28.3) | 81(38.2) | 32(15.1) | 39(18.4) | 55(26.2) | 82(39.0) | 37(17.6) | 36(17.1) |
| **CS is not necessary because a woman does not need to be cut before she delivers a baby** | 60(28.3) | 69(32.5) | 29(13.7) | 54(25.5) | 42(20.0) | 76(36.2) | 47(22.4) | 45(21.4) |

### Subjective Norms Disposition

The findings showed that subjective norms played a key role in shaping respondents’ acceptance of caesarean section (CS) delivery. Many respondents considered the opinions of their close social circles when deciding on CS. More than half of the male (59.4%) and female (59.5%) respondents would accept CS if their spouse supported it. Similarly, parental approval matters, with 68.4% of males and 66.7% of females more likely to accept CS if their parents viewed it as a good delivery method. Influence from in-laws is also significant, as nearly 7 in 10 males (69.4%) and 6 in 10 females (65.2%) would accept CS if their mother or father-in-law approved. Other family members also played a role, with 70.3% of males and 66.7% of females agreed that family support would make them more accepting of CS. Friends’ opinions affect acceptance as well, with 71.7% of males and 62.9% of females reported they would accept CS if their friends viewed it positively. However, healthcare professionals had slightly less influence, as only 58.5% of males and 51.4% of females would accept CS based on their doctor or nurse’s recommendation. Cultural and religious beliefs also shape acceptance of CS birth. About 68.9% of males and 64.2% of females reported CS has acceptable in their culture, also 64.6% of males and 63.3% of females reported that their religion permits it. Additionally, 67% of males and 64.7% of females would be more accepting of CS if they believed society viewed it as an acceptable method of delivery. Overall, these findings highlight the strong role of subjective norms influence in acceptance of CS with family, friends, and cultural beliefs playing a major role in shaping decisions. (refer to Table 3 for details).

**Table 3:**
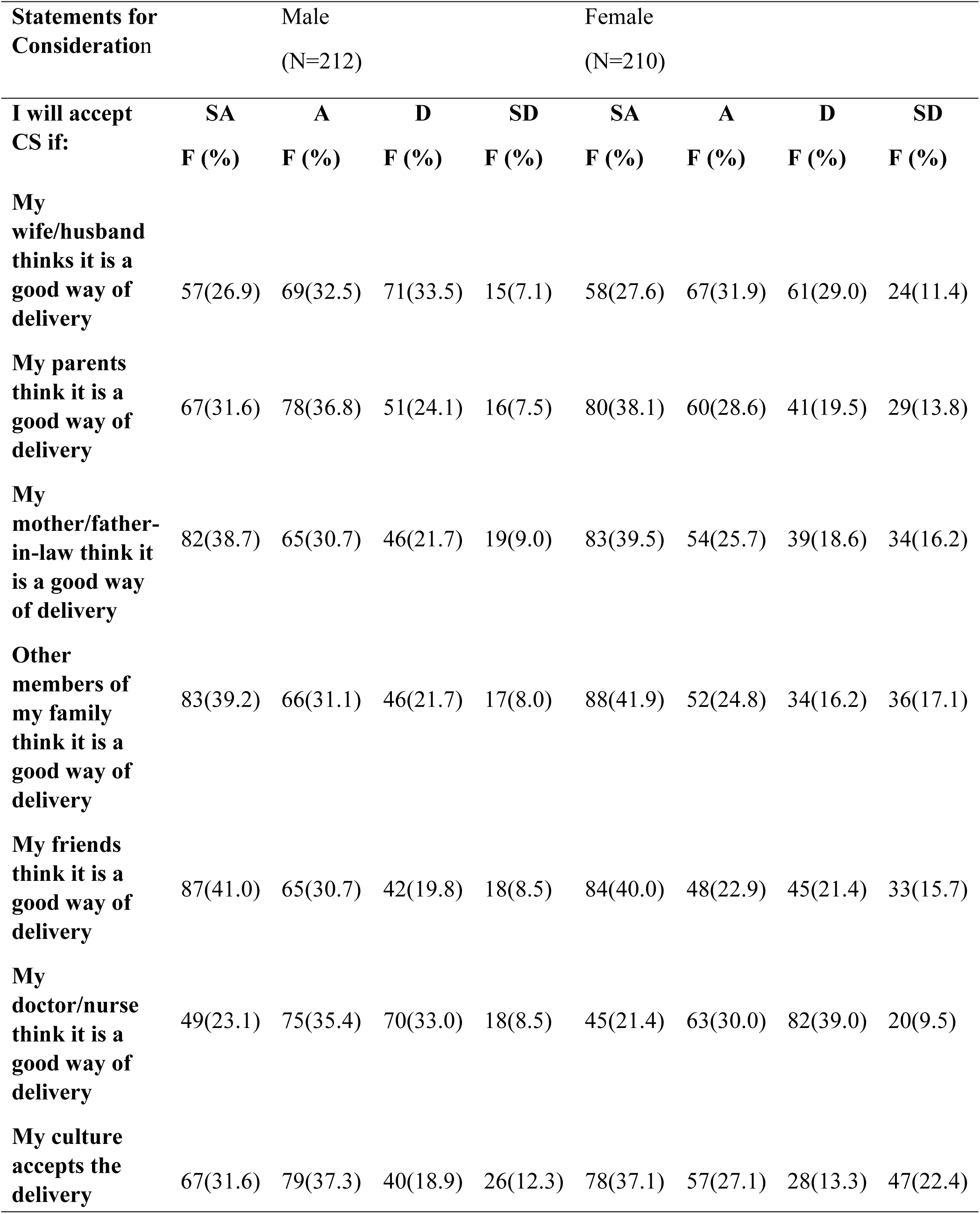
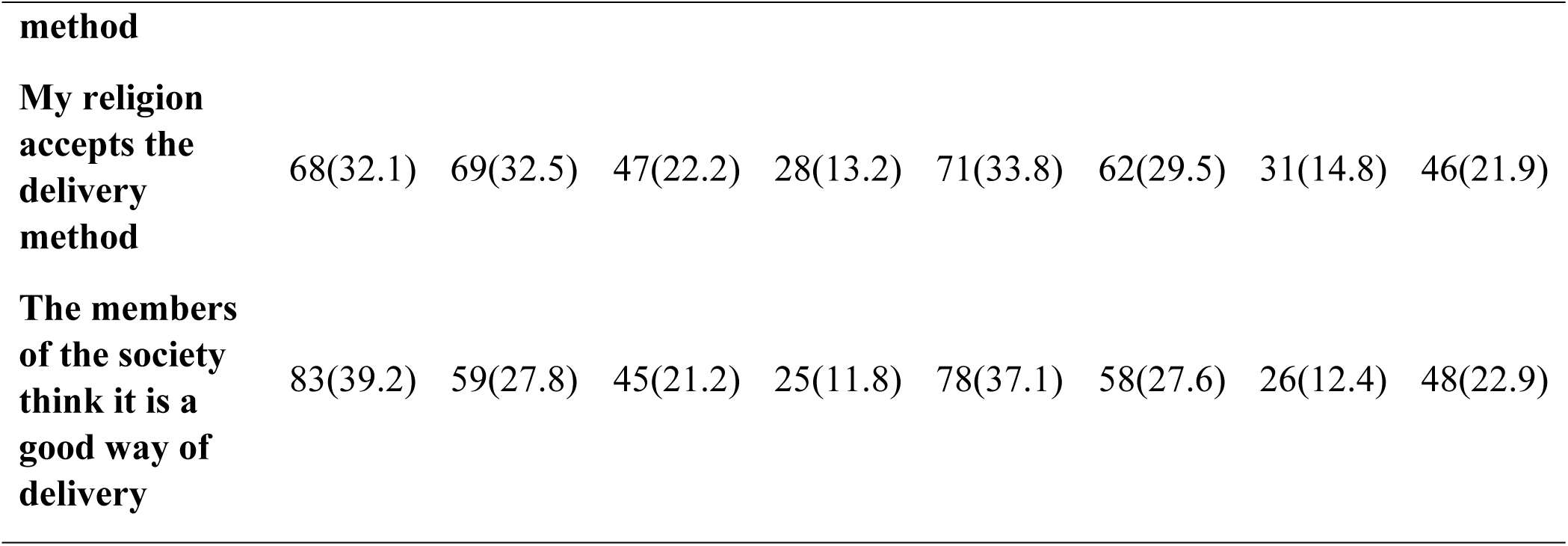
Findings on the Subjective norms influencing CS.

### Outcome Variable

#### Intention to Accept Caesarean Section (CS) Delivery

The frequency of responses on the intention to accept CS delivery in this study showed that only 3 out of every 10 male respondents and 2 out of every 10 females were likely to accept CS if the need arose. About 4 in 10 male and female respondents would accept CS as a less painful delivery method. Financial constraints influenced acceptance, as 56.1% males and 55.2% females were unlikely to opt for CS even if they could afford it. Similarly, 57.5% males and 55.2% females remained unwilling even if CS were free. Overall, CS acceptance remains high, with financial and personal factors influencing decisions (See Table 4 for full details).

**Table 4:**
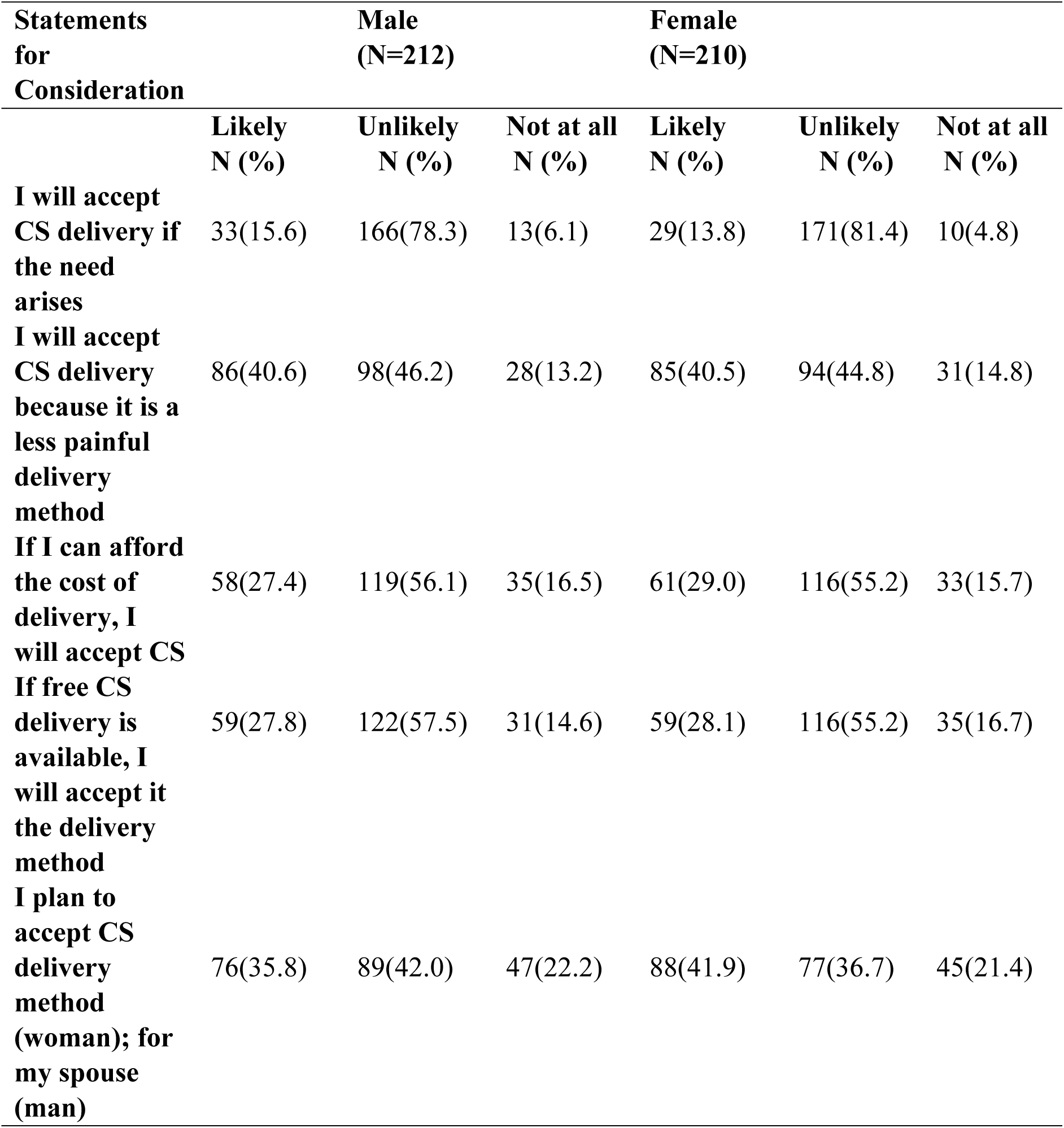
Findings on the intention to accept Caesarean Section (CS) delivery.

| **Statements**<br>**for**<br>**Consideration** | **Male**<br>**(N=212)** |  |  | **Female**<br>**(N=210)** |  |  |
| --- | --- | --- | --- | --- | --- | --- |
|  | **Likely**<br>**N (%)** | **Unlikely**<br>**N (%)** | **Not at all**<br>**N (%)** | **Likely**<br>**N (%)** | **Unlikely**<br>**N (%)** | **Not at all**<br>**N (%)** |
| **I will accept CS delivery if the need arises** | 33(15.6) | 166(78.3) | 13(6.1) | 29(13.8) | 171(81.4) | 10(4.8) |
| **I will accept CS delivery because it is a less painful delivery method** | 86(40.6) | 98(46.2) | 28(13.2) | 85(40.5) | 94(44.8) | 31(14.8) |
| **If I can afford the cost of delivery, I will accept CS** | 58(27.4) | 119(56.1) | 35(16.5) | 61(29.0) | 116(55.2) | 33(15.7) |
| **If free CS delivery is available, I will accept it the delivery method** | 59(27.8) | 122(57.5) | 31(14.6) | 59(28.1) | 116(55.2) | 35(16.7) |
| **I plan to accept CS delivery method (woman); for my spouse (man)** | 76(35.8) | 89(42.0) | 47(22.2) | 88(41.9) | 77(36.7) | 45(21.4) |

### Summaries of Descriptive Statistics for the Study Variables

#### Levels of TRA-constructs and Intention acceptances to CS

The findings displayed in Table 5 show the summaries of the statistics computed for the study variables descriptively. It can be observed that the level of attitudinal disposition among male respondents, measured on a 15-point maximum scale, is moderate (X-= 7.78) denoting 51.9% of the complete level expected from the respondents. The level of subjective norms (X-= 15.41) and intention (X-= 7.08) were equally reported with prevalence rates of 57.1% and 70.8%, respectively.

**Table 5:**
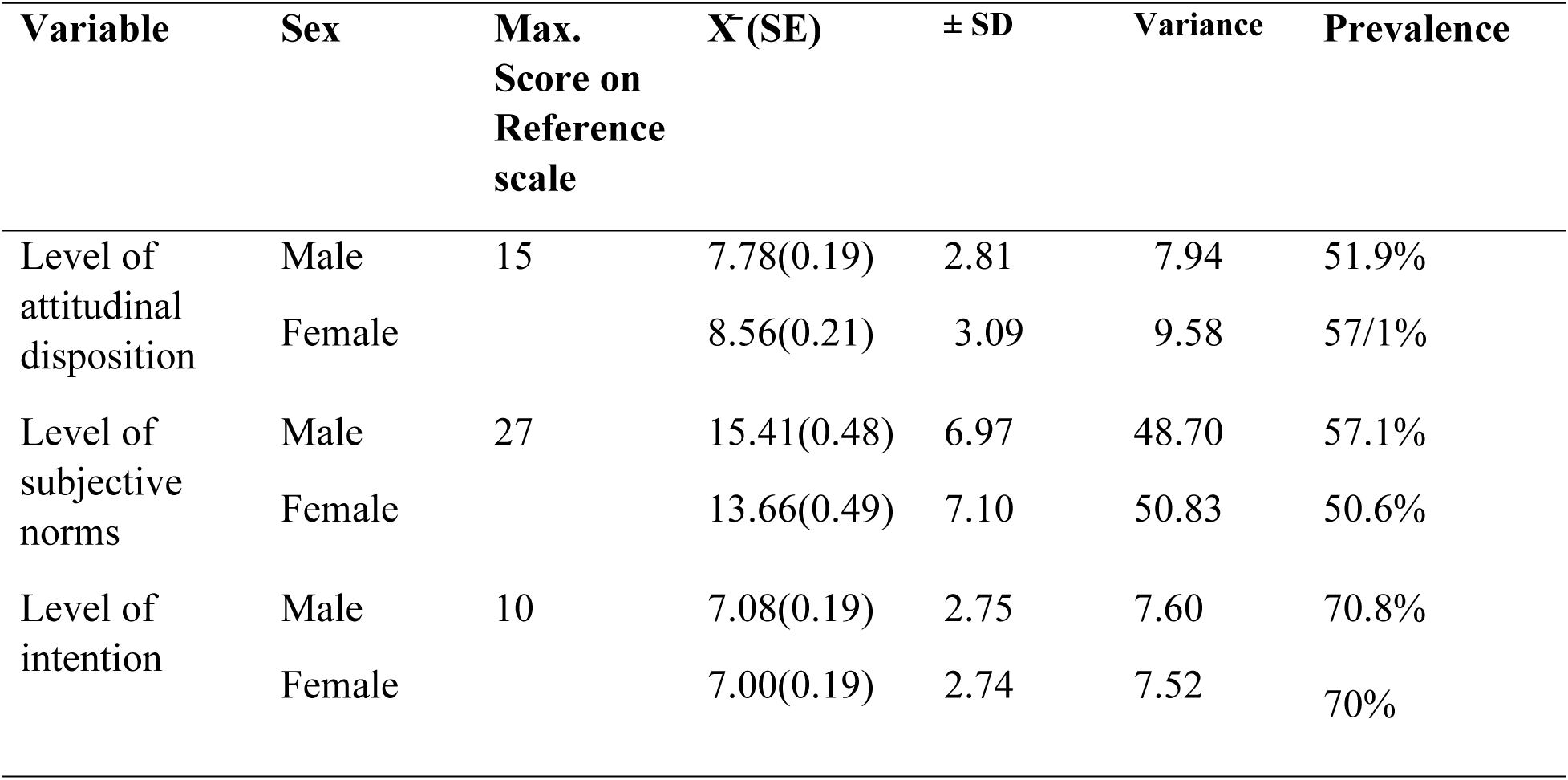
Summaries of descriptive statistics computed for the respondents.

| Variable | Sex | Max.<br>Score on<br>Reference<br>scale | $\bar{X}$ (SE) | ± SD | Variance | Prevalence |
| --- | --- | --- | --- | --- | --- | --- |
| Level of attitudinal disposition | Male | 15 | 7.78(0.19) | 2.81 | 7.94 | 51.9% |
|  | Female |  | 8.56(0.21) | 3.09 | 9.58 | 57/1% |
| Level of subjective norms | Male | 27 | 15.41(0.48) | 6.97 | 48.70 | 57.1% |
|  | Female |  | 13.66(0.49) | 7.10 | 50.83 | 50.6% |
| Level of intention | Male | 10 | 7.08(0.19) | 2.75 | 7.60 | 70.8% |
|  | Female |  | 7.00(0.19) | 2.74 | 7.52 | 70% |

Similarly, among female respondents, the level of attitudinal disposition (X-= 8.56) was slightly higher than that of males, denoting 57.1% of the complete level expected. The level of subjective norms (X-= 13.66) and intention (X-= 7.00) were also reported with prevalence rates of 50.6% and 70%, respectively (Refer to Table 5 for full details).

### Dynamics of CS Delivery Acceptance Based on Attitudinal Disposition and Subjective Norms

Table 6 presents the relationship between attitudinal disposition, subjective norm, and the intention to accept CS delivery. The R² values from the linear regression indicates that 4.4% and 9.5 % of the total change observed in the males and females intention to accept CS delivery can be explained by the TRA constructs. The F-value of 4.75 for males and 10.87 for females with a significant level of 0.002 and 0.001 respectively from analysis of variance indicated that the regression model predicts intention to accept CS delivery. Also the beta coefficient values (Males-B = 0.086, p = 0.002; Females-B = 0.121, p = 0.000) for subjective norm showed a significant positive relationship with intention to accept CS delivery (Refer to Table 6 for full details).

**Table 6:** Relationship between attitudinal disposition, subjective norms and Intention to accept caesarean section (CS) delivery.

| Sex | Variables | B | R<sup>2</sup> | F-value | p-value |
| --- | --- | --- | --- | --- | --- |
| Male | Attitudinal disposition | 0.061 | 0.044 | 4.75 | 0.382 |
| Female |  | 0.21 | 0.095 | 10.87 | 0.731 |
| Male | Subjective norm | 0.086 |  |  | 0.002 |
| Female |  | 0.121 |  |  | 0.000 |

## Discussion

This study assessed attitude and associated subjective norm involved in resolving whether to elect CS delivery among couples in Lagos, Nigeria.

### Attitude of respondents towards caesarean section delivery

This finding revealed that less than half of the respondents agreed that caesarean sections are an unnatural method of childbirth. Given that many respondents still believe vaginal delivery to be the best mode of childbirth; this finding corroborates previous findings where the respondents on the notion despite medical development^21^. Additionally, the respondents thought that having CS indicated that a woman was weak and unfaithful. A Study in sub-Saharan Africa also reported that women who had CS frequently experience mockery and were viewed as “less of a woman”^22^.

Regarding decision-making, less than half of the respondents affirmed spousal support for CS especially support of the husbands. This suggests that male dominance in reproductive health decisions is still common in many societies. In certain cultures, women’s participation in the informed consent process for CS is limited by cultural and gender norms, requiring consent from a male family member before the procedure can be performed^23^. Finding also revealed that the respondents perceived CS as a means for generating income for the doctors. This perception could contribute to mistrust of healthcare providers and may discourage pregnant women from seeking appropriate medical care. A study in Togo reported similar concerns, that pregnant women’s reluctance to undergo CS was partly due to mistrust in healthcare providers and fear of unnecessary medical interventions^23^.

In addition less than half of the respondents reported that CS is not necessary. This idea is the result of ignorance regarding delivery complications that require CS. A study in Ethiopia reported that women’s preference for CS was associated with factors such as dissatisfaction with previous intra-partum care and lack of knowledge about CS^24^.

### Subjective Norms influencing Caesarean section Acceptance

The finding of the study revealed that over a quarter of the respondents will accept CS delivery if their partner thought it was a good method of delivery. These findings are consistent with a recent study that highlights the importance of spousal support in decisions regarding delivery. Similarly, women who had supportive partners were more likely to accept CS^25^. The acceptability of CS is also significantly shaped by the involvement of parents. The finding of this study are also comparable to those of a study conducted by Elom et al.^26^ where mother and paternal figures frequently have an impact on decisions pertaining to birth, especially in situations involving extended families. Furthermore, in-laws—particularly mothers-in-law— played a significant influence on childbirth decisions in sub-Saharan Africa, supporting customs about delivery techniques ^26^.

In addition to spousal and parental support, friends and societal expectations also have a significant influence on the acceptability of CS. Rodriguez et al.^22^ reported that societal expectations and peer influence had a 64.7% rate of CS acceptance in Middle Eastern countries. Maitanmi et al^21^ found that social media influenced CS perceptions, with both positive and negative narratives influencing decision-making. The study also revealed that cultural and religious beliefs had a significant impact on CS acceptance. These findings are consistent with the findings of Sadore et al^27^, who pointed out that religious leaders have a significant influence on maternal health decisions in conservative societies. This could be because religious teaching combined with faith-based intervention to foster acceptance of CS among church and society members.

### Respondent Intention to Accept CS delivery

The majority of respondents stated that they were unlikely to accept CS delivery if the need arises. This could be brought on by mistrust of medical professionals, cultural beliefs, a fear of surgery, or a fear of dying. Similar obstacles to CS acceptance such as expense of the medical procedure fear of rejection from parents and husbands, fear of prejudice were also reported in a study in South-west Nigeria^28^. However, the finding revealed that more than half of the respondents were likely to accept CS if the service is free and affordable. The high cost associated with CS is a significant deterrent, as corroborated by studies highlighting economic constraints as a barrier to CS utilization^29^. Due to insufficient maternal education, financial constraints, and a lack of knowledge about Caesarean sections, less than half of the respondents—42.0% of the men and 41.9% of the women—remained reluctant to accept CS for their spouses. Adewuyi et al^29^ identified important factors that influence CS acceptance, including income index, maternal age, prenatal contact, birth order, birth type, birth size, frequency of internet use, maternal religion, and authorization to receive medical care. These factors are relevant to our findings. Therefore, there is need for a call to action on the urgent need to sensitize married couple, expectant mothers, youth and societies on the advantages and benefits of Caesarean section.

### Recommendations

It is important to launch comprehensive health education that will not only focused on pregnant women but it will also include their partners and in-laws during such programs efforts should be made to debunk misconceptions about CS, especially the idea that it is a sign of weakness or an unneeded medical procedure. Key stakeholders, such as religious and traditional leaders, should be involved in these efforts in order to encourage a change in social norms and encourage well-informed decision-making. The adoption of CS is still significantly hampered by financial constraints, which calls for government initiatives to increase access to free or reasonably priced CS services.

Further, increasing health insurance coverage and offering financial aid for CS in public hospitals will help reduce worries about costs and guarantee that those who need medical operations will be more likely to accept such method of delivery. Additionally, incorporating thorough CS education into antenatal care service will raise procedure awareness and acceptance. During antenatal visits, it is imperative that healthcare professionals provide individualized counselling that explains the advantages and medical justifications for CS in order to reduce fear, anxiety and misinformation, before the need arises. Public health initiatives can address the social and cultural factors impacting CS acceptance while also improving maternal and newborn health outcomes by putting these strategies into practice.

## Conclusion

This study reported a positive attitude toward caesarean section (CS), with high subjective norm considerations in accepting CS. Spouse, Family, friends, and cultural beliefs played a major role in decision-making, reinforcing societal expectations regarding childbirth delivery options. These factors, in turn, contributed to intention to accept CS delivery among the respondents in the study.

## Data Availability

Data has been provided as part of the submitted article

## Acknowledgements

we acknowledge all the research participants for their voluntary participation and cooperation throughout the data collection process.

## Authors Contribution

OT - conceptualized the study, discussion of findings, prepare the manuscript; AB-Literature review and data collection; ¶OB - Data collection and literature review; OO- literature review and proof read the manuscript; &PC Data collection, methodology; As; Data analysis and proof read the manuscript; EJ- Data collection and literature review; OA- Data analysis and Discussion of findings; AN- conceptual framework and proof read the manuscript.

